# Seroprevalence of SARS-CoV-2 and risk factors in Bantul Regency, Yogyakarta, Indonesia

**DOI:** 10.1101/2022.06.07.22276128

**Authors:** Riris Andono Ahmad, Citra Indriani, Risalia Reni Arisanti, Ratih Oktri Nanda, Yodi Mahendradhata, Tri Wibawa

## Abstract

COVID-19 case counts in Indonesia inevitably underestimate the true cumulative incidence of infection due to limited barriers to testing accessibility and asymptomatic infections. Therefore, community-based serological data are essential for understanding the true prevalence of infections. This study aims to estimate the seroprevalence of SARS-CoV-2 infection and factors related to the seropositivity in Bantul Regency, Yogyakarta, Indonesia. A cross-sectional study involving 425 individuals in 40 clusters was conducted between March and April 2021. Participants were interviewed using an e-questionnaire developed in the Kobo toolbox to collect information on socio-demographic, COVID-19 suggestive symptoms, history of COVID-19 diagnosis and COVID-19 vaccination status. A venous blood sample was collected from each participant and tested for immunoglobulin G (Ig-G) SARS-CoV-2 antibody titers using the enzyme-linked immunosorbent assay (ELISA). Seroprevalence was 31.1% in the Bantul Regency: 34.2% in semi-urban and 29.9% in urban villages. However, there is no significant proportion difference between both areas. A significant difference was reported in the age group. Participants in the 55-64 age group demonstrated the highest seroprevalence (43.7%; p=0.00), with a higher risk compared to the other age group (aOR= 3.79; 95% CI, 1.46-9.85, p<0.05). Seroprevalence in the unvaccinated participants was 29.9%. Family clusters accounted for 10.6% of the total seropositive cases. No significant difference was observed between individual preventive actions and their mobility with seropositivity status. This study observed a discrepancy with COVID-19 confirmed cumulative incidence data reported in the same period (11 out of 1000 population), indicating silent transmission may have occurred within the community. Higher seroprevalence in semi-urban areas rather than urban areas suggests a gap in health services access. Surveillance improvement through testing, tracing, and treatment, particularly in areas with lower access to health services, are necessary, along with more robust implementations of health protocols.

## Introduction

Coronavirus disease 2019 (COVID-19) is a respiratory illness caused by the newly discovered severe acute respiratory syndrome coronavirus 2 (SARS-CoV-2), leading to a global pandemic, including Indonesia. Since the first confirmed SARS-CoV-2 by Mar 2 2020 [1], Indonesia has experienced exponential growth of COVID-19 cases in 34 provinces. As of Mar 31 2021, the government of Indonesia reported a total of 1.511.712 confirmed COVID-19 cases and 40.858 deaths [2] and became one of the countries with high cumulative and incidence cases of COVID-19. However, these case counts inevitably underestimate the true cumulative incidence of infection because of limited diagnostic test availability, barriers to testing accessibility, and asymptomatic infections [2]. As a consequence, the national prevalence of SARS-CoV-2 remains unknown.

The lack of clarity about the number of SARS-CoV-2 infections across Indonesia limits the Indonesian government’s ability to plan appropriately, prepare and respond to this epidemic. Counting the incidence of newly diagnosed cases of severe COVID-19 and the case fatality rate is critical as it is necessary to project the need to address the demands on the healthcare system. The absence of a reasonable estimate of the number of infections poses a challenge for authorities to estimate this need.

One of the epidemiological investigations used to determine the level of disease spread is to apply a seroprevalence survey. According to the Centers for Disease Control and Prevention (CDC), this survey uses serological tests to detect antibodies in the blood, indicating an infection [3]. This test uses an enzyme-linked immunosorbent (ELISA) where the antigen used is purified SARS-CoV-2 S protein (without live virus). Population-based serological testing provides better estimates of the cumulative incidence of infection by complementing diagnostic testing of acute illness and helping to inform the public health response to COVID-19. A seroprevalence study could also be a powerful tool to detect subclinical infections and improve policy making in the country [4, 5]. Furthermore, as the world moves through the vaccine and variant era, synthesizing seroepidemiology findings is increasingly important to track the spread of infection, identify disproportionately affected groups, and measure progress towards herd immunity [2].

Seroprevalence varies geographically; the denser urban areas have higher seropositivity rates than rural areas [6]. A study in East Java, Indonesia, in the second semester of 2020 showed a higher prevalence in Surabaya (13.1%) which is an urban area, than in Jombang (9,9%), a rural area [7]. The epidemiological trend also implicates SARS-CoV-2 spread among rural communities only later in the epidemic wave [8, 9], which would require sound anticipatory interventions. A seroprevalence survey involving more diverse groups of people among urban and rural communities is necessary to grasp the overall picture of SARS-CoV-2 infection. Bantul regency has become one area that contributes to a high number of cases and leads to the high transmission of COVID-19 in Yogyakarta Provinces. Therefore, this study aims to estimate the SARS-CoV-2 prevalence, seropositive risk factors, and COVID-19 vaccine acceptance in Bantul Regency, Yogyakarta.

## Materials and methods

### Study setting

Bantul Regency is located in the southern region of Yogyakarta Province, covering 506.85 km^2^. The regency consists of 17 sub-districts and 75 villages, 30% semi-urban. The main occupation is the non-formal sector such as farmer, trade and services business [10]. The daily mobility of residents between districts to and from Bantul is high [11], which might risk disease transmission.

### Study design and sampling

A cross-sectional study was conducted from March to April 2021. The population in this study was individuals who lived in the Bantul regency for a minimum of six months, with a total of 1.018.402 inhabitants [10]. EpiInfo™ was used to calculate the sample size. The minimum sample size required was estimated based on a 20% of anti-SARS-CoV-2 antibodies prevalence, 6% sampling error, a significance level of 0.05 with a design effect of 2 and a non-response rate of 15%. Therefore, a total minimal sample size of 414 people.

Sampling was selected using a multistage cluster random sampling adopted from the WHO/EPI rapid survey [12]. Cluster size was 11, and clusters are neighbourhoods in a sub-village of around 50-70 households and selected using simple random sampling based on the elected village. A probability proportionate to size (PPS) sample was used to determine villages and clusters. We used a C-survey, 40 clusters distributed in 36 villages (25 urban and 11 semi-urban) and 17 subdistricts (48% from whole villages in Bantul). Systematic random sampling was used for recruiting households. Household members were defined as anyone who lived under one roof, ate from one kitchen, and resided in the study area for at least six months. In each household, a maximum of five eligible participants were recruited.

Participants must meet inclusion criteria as follows: residing in the study location at least six months before the survey commences, age of 5 years old or older, able to communicate verbally, give written consent to participate in the research or consent from parents/ guardians for respondents under 18 years old. Meanwhile, the exclusion criteria include people who cannot be sampled due to untoward illness (including immunocompromised individuals who have a history of blood disorders and people with mental disorders).

### Data collection

Door to door visit was conducted for collecting the primary data. Written informed consent was obtained from each study participant before data collection. Fifteen enumerators, including a phlebotomist, were involved in the data collection. Two supervisors were assigned to ensure the methodology and conduct the spot check. All study teams were involved in four training days regarding methods, data entry, phlebotomy and ethics in research.

Two milliliters of blood were drawn from a cubital vein and kept in an EDTA tube. Each participant’s data included a unique identifier (barcode label) linked to their blood sample and data for tracking and confidentiality. The blood sample was further transported, examined and stored in the Laboratory of Microbiology, Faculty of Medicine, Public Health, and Nursing UGM. Plasma was then tested for Ig-G anti-SARS-Cov2 procedures using (Human Anti-2019 n-CoV(N) IgG ELISA Kit V1.5 FineTest [13].

Risk factors information at household and individual levels were obtained using an electronic structured questionnaire developed in the KoBo toolbox. Variables included in the household questionnaire were socio-demographic information (age, gender, relationship with the head of household, and household income). Individual questionnaires observed socio-demographic information (age, gender, highest education, occupation), COVID-19 vaccination status, previous diagnosis of COVID-19 and symptoms related to COVID-19 within the last six months, preventive actions taken and mobility in the previous two weeks. The data manager validated data daily. A day after the data entry, the data manager sent the supervisor feedback and confirmation regarding completeness and data consistency.

Updated cumulative data on notified cases were obtained through the COVID-19 surveillance system conducted by Bantul District Health Office Bantul from March to April 2021. COVID-19 was defined as an asymptomatic or asymptomatic person with positive PCR results tested for SAR-CoV2. Population numbers per sub-district were obtained through Bantul Statistical Bureau for calculating incidence per 1000 population [10]. Data on urban-rural classification was obtained from the National Statistical Bureau [14].

### Statistical analysis

The seroprevalence and socio-demographic characteristics of study participants were described proportionally using the table. Map developed in ArcGIS was used to visualize the comparison between seroprevalence and incidence of notified cases by district and urban-rural status. Bivariate analysis was conducted to identify the association between presumed risk factors (age, gender, occupation, comorbidities, prevention taken and mobility over the last two weeks) and anti-SARS CoV-2 seropositivity. After considering collinearity and interaction, a multiple logistic regression was used to examine the relationship between the independent variable and Anti-SARS CoV-2 seropositivity while controlling for the effects of other covariates. The P-value would be considered statistically significant at p < 0.05. All statistical analysis was performed using STATA 14.0.

### Ethics

The study was approved by the Medical and Health Research Ethics Committee of the Faculty of Medicine, Public Health and Nursing Universitas Gadjah Mada with the number KE/1242/12/2020. Written informed consent was obtained from adult respondents and parents of enrolled children. Confidentiality of information from the respondents was upheld with utmost care throughout data collection, processing and analysis for all data collected. Therefore, their names were included in the notes only for traceability and referral during the data analysis.

## Result

### Characteristics of study participants

The prevalence of SARS-CoV-2 seropositivity among the participants in this study was 31.1% (n=132/425). A significant difference was observed in the seroprevalence among age groups (p=0.000), with the highest proportion reported in the 55-64 years age group (43.7%; n=31/71). Meanwhile, the under 15 years age group showed no seropositivity.

A significant difference was also observed among occupation groups (p=0.009). The highest seroprevalence was demonstrated by participants working as daily workers/farmers (37.2%), followed by professional/health workers (34.6%), and unemployed/students/housewives (26%). Seroprevalence does not differ between semi-urban and urban areas even though we observed that semi-urban areas have higher seroprevalence (34.2%; n=40/117) than urban areas (29.9%; n=92/308). In terms of gender, we found no significant differences in prevalence between males and females, even where females showed higher seropositivity than males (Table 1).

**Table 1.** Distribution of Study Participants by Background Characteristics.

| Characteristic | Total | IgG-positive | IgG-negative | P-value |
| --- | --- | --- | --- | --- |
|  | n = 425 | n=132 | n=293 |  |
| Sex, n (%) |  |  |  |  |
| Male | 174 (40.9) | 52 (29.9) | 122 (70.1) | 0.663 |
| Female | 251 (59.1) | 80 (31.9) | 171 (68.1) |  |
| Age group, years, n (%) |  |  |  |  |
| <=14 | 16 (3.8) | 0 (0.0) | 16 (100) |  |
| 15-24 | 52 (12.2) | 8 (15.4) | 44(84.6) |  |
| 25-34 | 51 (12.0) | 8 (15.7) | 43(84.3) |  |
| 35-44 | 90 (21.2) | 33 (36.7) | 57(63.3) | 0.000 |
| 45-54 | 92 (21.7) | 32 (34.8) | 60 (65.2) |  |
| 55-64 | 71 (16.7) | 31 (43.7) | 40 (56.3) |  |
| 65+ | 53 (12.5) | 20 (37.7) | 33 (62.3) |  |
| Occupation, n (%) |  |  |  |  |
| Unemployed/students/housewife | 200 (47.2) | 52 (26.0) | 148 (74.0) | 0.009 |
| Professional/health worker | 130 (30.7) | 45 (34.6) | 85 (65.3) |  |
| Daily worker/farmer | 94 (22.2) | 35 (37.2) | 59 (62.8) |  |
| Residence setting, n (%) |  |  |  |  |
| Urban | 308 (72.5) | 92 (29.9) | 216 (70.1) | 0.390 |
| Semi-urban area | 117 (27.5) | 40 (34.2) | 77 (65.8) |  |
| Smoking, n (%) |  |  |  |  |
| Smoker | 74 (18.1) | 14 (18.9) | 60(81.1) | 0.007 |
| Non smoker | 335 (81.9) | 118 (35.2) | 217(64.8) |  |
| History of chronic disease, n (%) |  |  |  |  |
| Yes | 96 (22.6) | 37 (38.5) | 59 (61.5) | 0.072 |
| No | 329 (77.4) | 95 (28.9) | 234 (71.1) |  |
| Diabetes mellitus, n (%) |  |  |  |  |
| Yes | 15 (3.5) | 6 (40.0) | 9 (60.0) | 0.514 |
| No | 394 (92.5) | 126 (32.0) | 268 (68.0) |  |
| Hypertension, n (%) |  |  |  |  |
| Yes | 65 (15.9) | 23 (35.4) | 42 (64.6) | 0.559 |
| No | 344 (84.1) | 109 (31.7) | 235 (68.1) |  |
| Obesity, n (%) |  |  |  |  |
| Yes | 8 (1.9) | 4 (50) | 4 (50) | 0.279 |
| No | 401 (98.1) | 128 (31.9) | 273 (68.1) |  |
| Previous COVID-19 diagnosis, n (%) |  |  |  |  |
| Yes | 3 (0.7) | 1 (33.3) | 2 (66.7) | 1.000 |
| No | 422 (99.3) | 131 (31.0) | 291 (68.9) |  |
| COVID-19 symptoms n (%) |  |  |  |  |
| Yes | 155 (36.4) | 44 (28.4) | 111 (71.6) | 0.4 |
| No | 270 (63.4) | 88 (32.6) | 182 (67.4) |  |

| COVID-19 vaccination, n (%) |  |  |  |  |
| --- | --- | --- | --- | --- |
| Yes, at least 1 dosage | 13 (3.1) | 9 (69.2) | 4 (30.8) | 0.005 |
| Not yet | 412 (96.9) | 123 (29.9) | 289 (70.2) |  |

This study also explored the seroprevalence based on chronic diseases and comorbidities history. Participants with a history of chronic conditions accounted for 22.6% of the total participants, and comorbidities identified in this study were diabetes mellitus, hypertension and obesity. Among those with comorbidities, seroprevalence for participants with obesity, diabetes mellitus, and hypertension were 50%, 40%, and 35.4%, respectively. Participants with a prior diagnosis of COVID-19 in the last six months accounted for 1 out of 132 seropositive cases. Seroprevalence among participants with no previous experience of COVID-19 related symptoms was found to be 31.1%. Seropositivity was observed in 14 participants under seven different households, making family clusters account for 10.6% of the total positive cases.

In terms of preventive actions, seroprevalence was found highest in participants who reported not always taking the preventive measures such as 1) wearing masks when going out (35.6%); 2) maintaining a physical distancing (1-2 m) in the public area (33.1%); and 3) washing hands for at least 20 seconds (32.1%). Meanwhile, in the mobility aspect, the highest seroprevalence was observed in participants who always attended invitations to traditional or religious activities (35.9%). Frequent mobility in visiting relatives/friends for essential matters and going to the shops/markets/offices contributed to 30.9% and 30.1% of seroprevalence, respectively. Of the participants who reported not always staying at home, the seroprevalence was 33.9%. No significant difference was observed between individual preventive actions and their mobility with seropositivity status (Table 2).

**Table 2.** Distribution of Study Participants by Preventive Action and Mobility.

| Preventive measure and mobility | Total | IgG-positive | IgG-negative | p-value |
| --- | --- | --- | --- | --- |
|  | n = 425 | n=132 | n=293 |  |
| Preventive measure |  |  |  |  |
| Wearing masks when going out |  |  |  |  |
| Always | 380 (89.4) | 116 (30.5) | 264 (69.5) | 0.491 |
| Not always | 45 (10.6) | 16 (35.6) | 29 (64.4) |  |
| Washing hands for at least 20 seconds with running water |  |  |  |  |
| Always | 344 (80.9) | 106 (30.8) | 238 (69.2) | 0.822 |
| Not always | 81 (19.1) | 26 (32.1) | 55 (67.9) |  |
| Maintain a physical distancing (1-2m) in the public area |  |  |  |  |
| Always | 274 (64.5) | 82 (29.9) | 192 (70.1) | 0.497 |
| Not always | 151 (35.5) | 50 (33.1) | 101 (66.9) |  |
| Mobility |  |  |  |  |
| Attending invitations to traditional or religious activities (e.g. weddings, funerals) |  |  |  |  |
| Always | 39 (9.2) | 14 (35.9) | 25 (64.5) | 0.493 |
| Not always | 386 (90.8) | 118 (30.6) | 268 (69.4) |  |
| Visiting relatives or friends or other people due to important matters |  |  |  |  |
| Always | 42 (9.9) | 13 (30.9) | 29 (69.1) | 0.987 |
| Not always | 383 (90.1) | 119 (31.1) | 264 (68.9) |  |
| Going to the markets/shops/offices/crowds |  |  |  |  |
| Always | 166 (39.1) | 50 (30.1) | 116 (69.9) | 0.738 |
| Not always | 259 (60.9) | 82 (31.7) | 177 (68.3) |  |
| Staying at home, except for essential matters |  |  |  |  |
| Always | 313 (73.7) | 94 (30.1) | 219 (69.9) | 0.444 |
| Not always | 112 (26.3) | 38 (33.9) | 74 (66.1) |  |

### Seroprevalence of anti-SARS-CoV-2 based on the geographical distribution

This study reported no difference between urban and semi-urban areas. However, we observed higher seropositive cases in semi-urban areas, of which higher numbers were found in Pajangan (56.5%), Dlingo (42.1%), and Sanden (41.7%) (Figure 1). The highest distribution of confirmed cases acquired from routine regional data reported a higher cumulative cases number in urban areas, such as Banguntapan, Bantul, Sewon, and Jetis.

**Fig 1.**
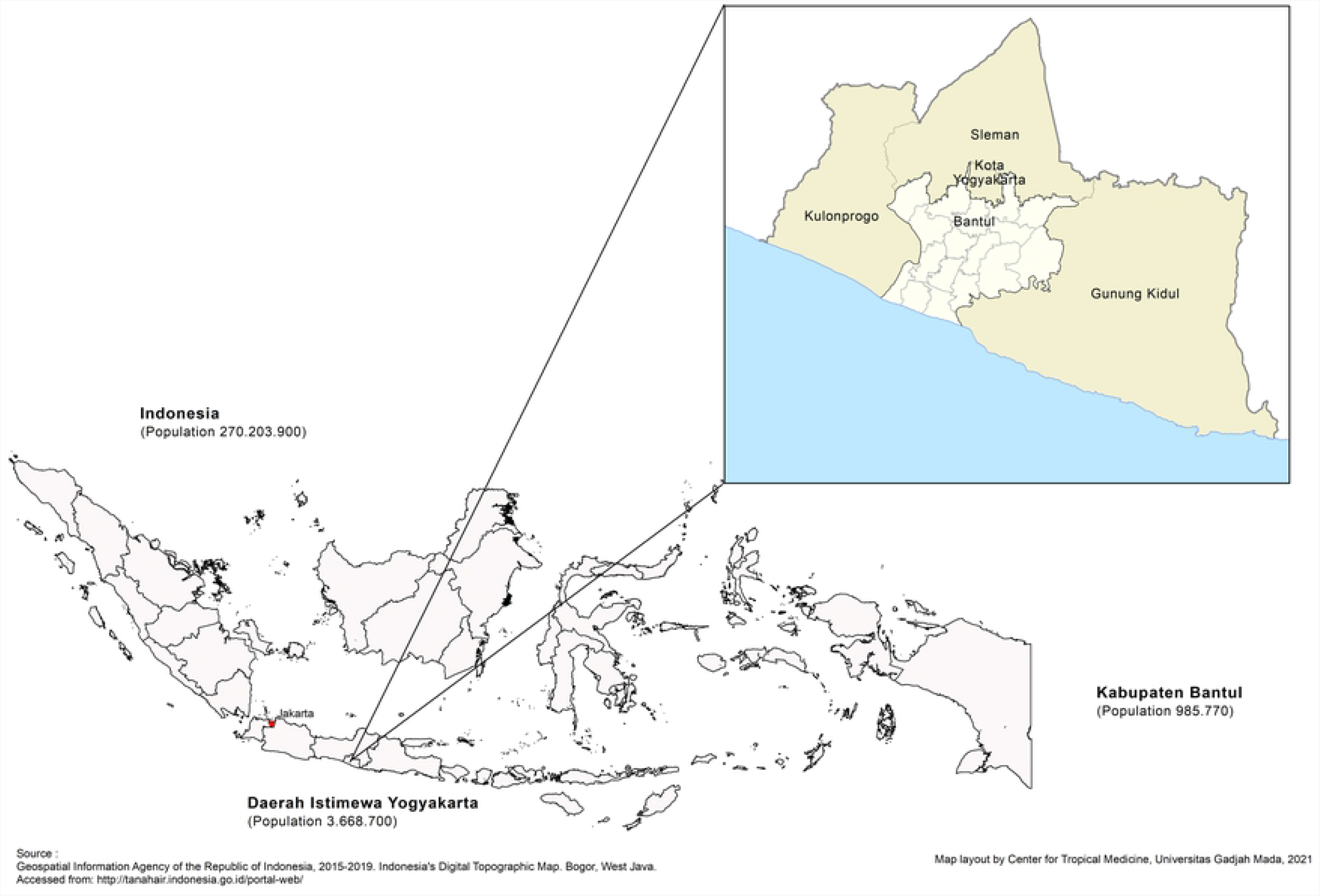
Map of Bantul Regency, Special Region of Yogyakarta, Indonesia.

**Fig 2.**
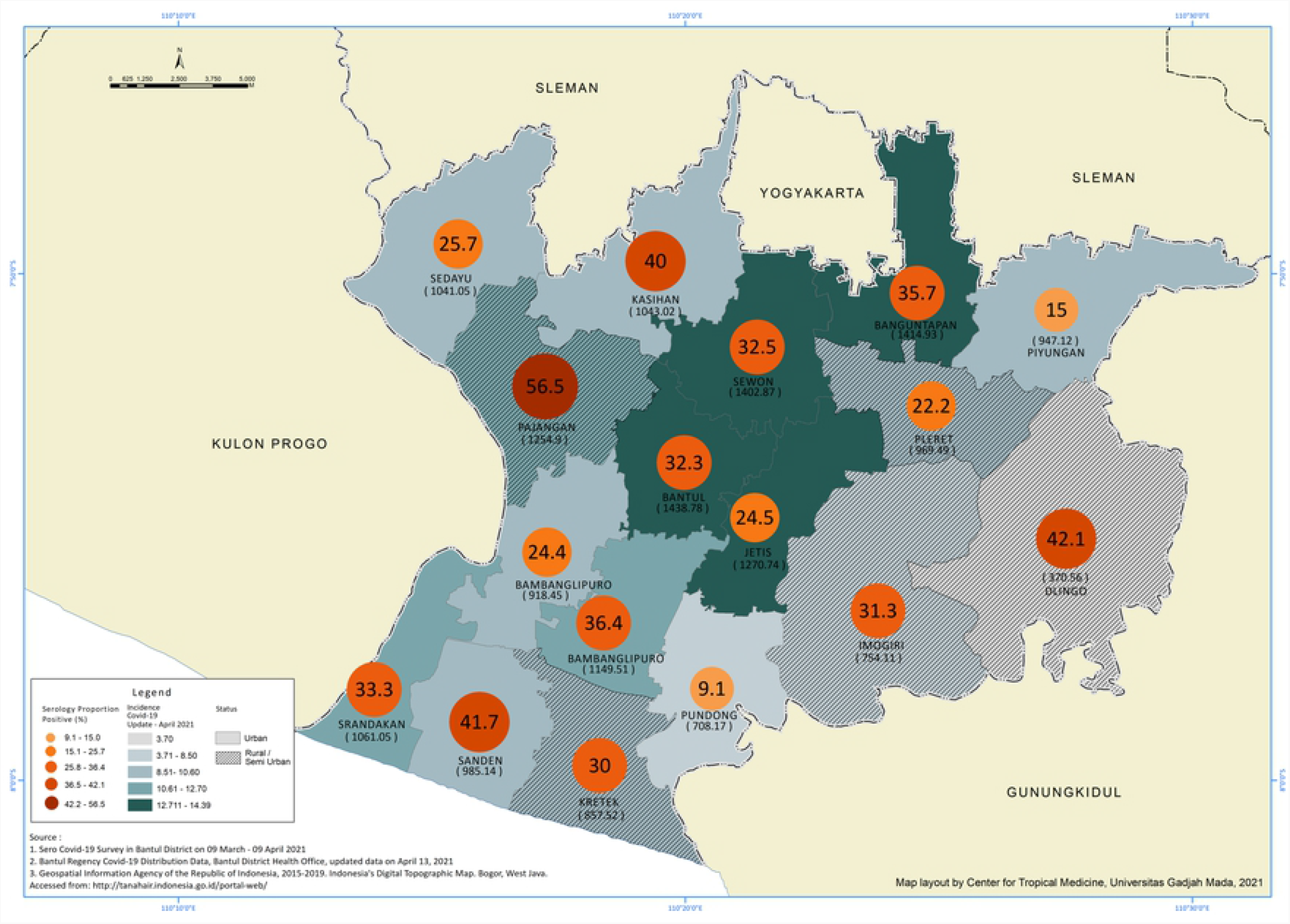
Seroprevalence and Reported Cumulative Incidence of SARS-Cov-2 in April 2021 Based on District.

### Risk factors associated with SARS-CoV-2 seropositivity

This study further explored the risk factors associated with the seropositivity of SARS-CoV-2, adjusting for age, sex, occupation, comorbidities, and vaccination status. A significant association was observed within the age group. The odds of SARS-CoV-2 seropositivity is higher in the age 55-64 (adjusted odds ratio [aOR]=3.79; 95% CI 1.46-9.85, p=0.006). Although females demonstrated higher seroprevalence, no association was found between gender difference and occupation. (aOR=0.81; 95% CI 0.50-1.31, p>0.005) (Table 3).

**Table 3.** Factors Associated with SARS-Cov-2 Seropositivity in Bantul Regency (N=425).

| **Risk Factors** | **ELISA+ (n/N)** | **Adjusted OR (95% CI)** | **p-value** |
| --- | --- | --- | --- |
| **Age group** |  |  |  |
| <=14 | 0/132 | NA | NA |
| 15-24 | 8/132 | Reference | NA |
| 25-34 | 8/132 | 0.93 (0.31-2.79) | 0.893 |
| 35-44 | 33/132 | 2.71 (1.08-6.81) | 0.034 |
| 45-54 | 32/132 | 2.67 (1.05-6.80) | 0.040 |
| 55-64 | 31/132 | 3.79 (1.46-9.85) | 0.006 |
| 65+ | 20/132 | 3.38 (1.27-9.00) | 0.015 |
| **Sex** |  |  |  |
| Male | 52/132 | 0.81 (0.50-1.31) | 0.388 |
| Female | 80/132 | Reference | NA |
| **Occupation** |  |  |  |
| Unemployed/students/housewife | 52/132 | Reference | NA |
| Professional/health worker | 45/132 | 1.14 (0.64-2.00) | 0.661 |
| Daily worker/farmer | 35/132 | 1.27 (0.70-2.31) | 0.441 |
| **Comorbidities** |  |  |  |
| Hypertension | 23/132 | 0.87 (0.48-1.57) | 0.646 |
| Obesity | 4/132 | 2.46 (0.57-10.65) | 0.229 |
| Diabetes Mellitus | 6/132 | 1.12 (0.38-3.31) | 0.832 |
| **Vaccination Status** |  |  |  |
| Yes | 14/132 | Reference | NA |
| No | 13/132 | 0.19 (0.05-0.68) | 0.010 |

Comorbidities were presumed as one of the risk factors of SARS-CoV-2 seropositivity. The odds of SARS-CoV-2 seropositivity are higher in participants with obesity (aOR=2.46; 95% CI 0.57-10.65) and diabetes mellitus (aOR=1.12; 95% CI 0.38-3.31) than participants without each respective comorbidity. However, this finding is not statistically significant (p-value>0.05).

Although vaccination has not been rolled out to the general public during the data collection, the majority of the participants (91.5%) were aware that the government would provide the vaccination for the people and stated their willingness (79.1%) to get vaccinated. Only a small proportion of the participants (11.1%) refused to get vaccinated for various reasons, such as being afraid of the adverse effect, uncertain safety and effectiveness, and religious beliefs.

## Discussion

The finding from this seroprevalence study for SARS-CoV-2 reported that the prevalence of IgG antibodies against SARS-CoV-2 was 31.1% in Bantul when the vaccination program had just started targeted health workers. It indicates higher seropositivity than 11.4% in a study in East Java in late 2020 [7]. The discrepancy between our research and COVID-19 cumulative incidence notified cases in the Bantul Regency, which reported a 1.1% cumulative incidence within the same period. This finding is in line with previous research that suggested the estimates obtained from seroprevalence were 18.1 times higher than the corresponding cumulative incidence of COVID-19 infections, implying that confirmed cases as a poor indicator of the extent of the disease spread [15].

Seroprevalence varies geographically, and previous research indicated that urban areas mainly reported a higher number of seropositivity [6]. Regional data reported a higher prevalence in urban areas in Bantul (Bantul, Banguntapan, Sewon, and Jetis). However, this study showed that higher seropositivity was descriptively observed in semi-urban areas (Pajangan, Dlingo and Sanden), suggesting a disparity in access to health services.

At the pandemic’s beginning, urban areas with dense populations were impacted earlier. A previous study on epidemiological modelling predicted that COVID-19 spread faster in urban than rural areas [16]. Despite the rapid transmission, urban areas are usually supported by better surveillance management, such as more accessible testing, tracing, and control of the infection. Consequently, cases in urban areas were reported faster and earlier.

Meanwhile, rural areas usually have shortages in resources and workforces, which affects their ability to detect, respond, prevent, and control infectious disease outbreaks [17, 18]. In this study, a large proportion of seropositivity was reported in participants with no prior diagnosis of COVID-19, indicating that the infection might be undiagnosed due to a lack of individual testing. Thus, this study suggests that the higher transmission observed in semi-urban areas was lower access to health services, including lack of individual testing, resulting in unreported positive cases.

Higher seroprevalence in this study was observed in females. However, no statistically significant gender difference was reported. A previous study also suggested that SARS-CoV-2 IgG/IgM dynamic is mainly affected by age and disease severity, not sex [19]. Higher seropositivity was observed in the age group 55-64, and this finding supports many previous studies that indicate older age as one of the risk factors for COVID-19. Though not statistically significant, this study found a higher risk of infection in participants with obesity. As suggested in a previous study, obesity now emerges as an essential risk factor for COVID-19 with more severe symptoms and an adverse prognosis [20].

This study subsequently found that a proportion of seropositivity was present among those with a frequent stay at home except for essential matters. In addition, occupation seemingly requires lower mobility, such as unemployed, students, and housewives, also accounted for seropositivity cases. As this study found one-tenth of family clusters, these findings indicate the transmission might also have occurred at the household level. Previous studies suggest that household transmission is considered a dominant route of COVID-19 infection [21, 22].

Household transmission might have occurred due to the challenging and ineffective separation and quarantine for higher mobility individuals in the same household [23]. The transmission at the household level may further spread to the community by those individuals. In this study, occupations requiring higher mobility, for instance, daily workers, farmers, and professional/health workers, have a higher seroprevalence. This study suggests that frequent mobility and lower compliance to preventive measures (wearing masks, washing hands, and physical distancing) also led to a higher number of seropositivity.

Surveillance and containment measures such as large-scale social restrictions and other anticipatory prevention become priority to curb the transmission, primarily focusing on areas with lower access to health services. Earlier in the pandemic, outbreak containment was concentrated in the dense urban areas, which may have reduced the number of cases. However, the transmission slowly moved to the semi-urban and rural communities due to high mobility. People living in rural communities might develop a false sense of security and take fewer precautions at the beginning than the urban communities [8,9].

One of the effective ways to prevent the fatalities caused by COVID-19 is through vaccination. Due to no vaccination programme rolled out to the general population during the data collection period, it is reasonable that higher seroprevalence was found in unvaccinated groups. As this study explored the seroprevalence when most people were still unvaccinated, a better picture of the infection spread in Bantul was obtained since there was no implication from vaccine-induced antibodies.

Despite the findings, this study is not without limitations. Following a natural infection, antibody titers peak and begin to wane in a various manner. This study, however, did not take into account the assay performance in relation to the waning immunity. Furthermore, as a cross-sectional study, this study only analyzed the variables at once and did not explore the seroprevalence changes over time as obtained in longitudinal studies.

## Conclusion

This serosurvey demonstrated a higher seroprevalence compared to reported data in the same period. Based on the findings, it is strongly recommended for the local government to strengthen the surveillance and 3T (testing, tracing, and treatment) efforts by involving the task force at the neighbourhood community and village levels throughout Bantul Regency, particularly in areas with lower access to health services. Besides that, increase awareness and implementation of health protocols, especially when carrying out activities with high mobility, to prevent transmission within the household. As the vaccination for the general population is being rolled out, it is crucial to provide adequate implementation information, including health resources and logistics. This study can be implemented in other areas, both at the district/city and provincial levels, to better understand the seroprevalence of SARS-CoV-2 in Indonesia. This study provides a district-level view of the extent of COVID-19 spread and a different approach to conducting serosurvey among diverse populations in various regions to fit the gaps in understanding COVID-19 spread in a global level.

## Data Availability

The data that support the findings of this study are available from the corresponding author upon reasonable request.

## Acknowledgments

We thank all study participants involved in the study, Bantul DHO, for supporting the research and all of the enumerators and the supervisor for the great work during the data collection.

## Supporting information

**S1 Fig. Map of Bantul Regency, Special Region of Yogyakarta, Indonesia**.

**S2 Fig. Seroprevalence and Reported Cumulative Incidence of SARS-Cov-2 in April 2021 Based on District**.

**S1 Table. Distribution of Study Participants by Background Characteristics**.

**S2 Table. Distribution of Study Participants by Preventive Action and Mobility**.

**S3 Table. Factors Associated with SARS-Cov-2 Seropositivity in Bantul Regency (N=425)**.

